# Association Between Uric Acid and Hemorrhagic Transformation: A Systematic Review and Meta-Analysis

**DOI:** 10.1101/2023.12.17.23300115

**Authors:** Ying Qian, Na Li, Yuanyuan Li, Chenxi Tao, Zhenhong Liu, Guoxia Zhang, Fan Yang, Hongrui Zhang, Yonghong Gao

## Abstract

**Background:** The relationship between uric acid (UA) and hemorrhagic transformation (HT) remained controversial. The purpose of this study was to investigate the relationship between UA degrees and the risk of HT after acute ischemic stroke (AIS).

**Methods and methods:** Electronic databases were sought for studies on UA and HT from inception to October 31, 2023. Two researchers independently reviewed the studies for inclusion. STATA Software 16.0 was used to compute the standardized mean difference (SMD) and 95% confidence interval (CI) of the pooled and post-outlier outcomes. The heterogeneity was evaluated using the I^2^ statistic and the Galbraith plot, and sensitivity analysis was also performed. Begg’s funnel plot and Egger’s test were used to assess publication bias.

**Results:** 12 trials were included in the meta-analysis, with a total of 4,708 individuals. Low UA degrees were linked to higher HT post-AIS patients following the pooled SMDs’ forest plot (SMD = -0.34, 95% CI = -0.60–0.08, *P* = 0.010). The high heterogeneity (I^2^ = 89.0%, *P*<0.001) was found in the studies. Six papers were outside the Galbraith plot regression line and there was no heterogeneity when they were excluded (I^2^ = 45.8%, *P* = 0.100). Meanwhile, the repeated SMDs (SMD = -0.487, 95% CI = -0.632–0.342, *P* = 0.000) still meant that the HT group had lower UA degrees. There was no publication bias in our meta-analysis following Begg’s funnel plot or Egger’s test.

**Conclusions:** The meta-analysis illustrated a substantial positive connection between UA degrees and HT, with lower UA separately linked with higher HT post-AIS. It provided a certain theoretical basis for the later related research.

## Introduction

Acute ischemic stroke (AIS) is a leading reason of the primary contributor and mortality to long-term handicap on a global scale^1–3^. This condition is effectively manageable by using thrombolytic therapy (IVT) or endovascular thrombectomy (EVT)^4, 5^. Hemorrhagic transformation^6^ (HT) can happen either as a natural progression of AIS or as a complication that results from AIS treatment. Severe neurological deterioration can be caused by HT, and even minor instances of HT can adversely affect long-term functional outcomes. As a result, it is vital to further our apprehension of HT to enhance the prognosis for AIS patients.

As reported by the Heidelberg criteria and the European Cooperative Acute Stroke Study classification^3^, HT can be classified as either parenchymal hematoma (PH) or hemorrhagic infarction (HI); it can likewise be divided into asymptomatic intracranial hemorrhage (ICH) and symptomatic ICH (sICH), depending upon whether neurological deficits are present or absent. Ample hazard factors^6–8^ that were associated with the event of HT have been identified, including age, the stroke’s severity, hyperglycemia, hypertension, the grade of blood calcium and uric acid, and the degree of cholesterol. The increase of blood-brain barrier (BBB) permeability caused by the inflammatory process of free radical release is one of the main causes of HT^9^.

Uric acid (UA) is derived from the metabolism of purines^3, 10^. Previous studies^11–13^ have illustrated that excessive UA degrees are linked to a raised risk of several disorders, including hypertension, chronic renal disease, cardiovascular and cerebrovascular diseases, etc. As stated by the studies^14, 15^, UA degrees seemed to be a separate hazard factor for dying early in AIS patients. UA, on the other hand, is the most substantial endogenous antioxidant^16^, as reported by an increasing number of studies. It exerts neuroprotective benefits by scavenging free radicals, preventing the inflammatory cascade, and reducing BBB permeability. For instance, investigations by Lei et al.^17^ and Wang et al.^10^ discovered that the amount of UA may possess a protective influence on the neurological outcome post-AIS. Therefore, any potential correlation between UA and AIS prognosis is unclear. At present, many studies have focused on the relationship between UA degrees and HT. But unfortunately, there was no agreement regarding the controversial role of US in HT after AIS. Thus, in the current investigation, a meta-analysis was conducted to evaluate the correlation between UA degrees and HT post-AIS.

## Methods

This meta-analysis was performed and reported following a predefined protocol (PROSPERO registration number: CRD####) (Registered but no results yet)and the Preferred Reporting Items for Systematic Reviews and Meta-Analyses (PRISMA) guidelines.

### Search strategy

Major databases, like PubMed, Cochrane Library, Embase, Web of Science, CBM (Chinese Biomedical Literature Database), CNKI (China National Knowledge Infrastructure), and Wanfang database, were thoroughly and meticulously searched for pertinent research. The search time was from inception to October 31,2023. The search terms were (“uric acid” OR “UA” OR “urate” OR “hyperuricemia”) AND (“Acute Ischemic Stroke” OR “AIS” OR “Ischemic Stroke” OR(“Stroke”) AND (“hemorrhagic transformation” OR ”HT”). Any additional articles identified were searched to expand the search. As all of the data we invoked were secondary summary data, no moral recognition was required for our study.

### Inclusion and exclusion criteria

The studies were analyzed by two reviewers (Ying Qian and Na Li) individually. Conflicts were resolved by consensus or with a superior investigator’s help (Yonghong Gao). Inclusion criteria: (1) full-text in Chinese or English; (2) AIS patients; (3) define inclusion criteria for AIS and HT; (4) comparison of UA degrees in HT and non-HT patients; (5) studies that reported UA degrees as a continuous variable or categorical variable (≥3 equal categories); (6) inclusion of only human participants. Exclusion Criteria: (1) incomplete information that cannot be extracted from the data literature; (2) conference abstracts, animal experiments, letters, comments, reviews, and case reports; and (3) while data from the same population was offered in multiple studies, a higher-quality study or a larger sample capacity was exclusively taken into account.

### Data extraction and quality assessment

All information was independently extracted by two investigators (Ying Qian and Na Li) using a standardized data collection form. Disagreements were resolved by a senior investigator (Yonghong Gao). From each included article, the following data was abstracted: the first author’s name, year, country, language, study design, number of patients, source of patients, mean age, gender, treatment, HT types, and time of assessment; the number of HT and non-HT patients; the UA degrees (mean and standard deviation, or median and interquartile range (IQR)) of patients; adjusted confounding factors; and conclusions. The Newcastle-Ottawa Scale (NOS)^18^ was used as the basis for carrying out the quality assessment. As per the NOS, papers with a grade of 9 stars were considered to be of the highest quality, while those with a grade of no less than 6 stars were considered to be of excellent quality.

### Statistical analysis

The STATA application (version 16.0, Stata Corp., College Station, TX) was used to analyze the abstracted information. UA degrees reported as median and IQR were converted to mean ±SD^19, 20^. Since the dates used were expressed as mean ±SD, the standardized mean difference (SMD) and 95% confidence interval (CI) were computed for each study. After that, the computed result was combined and checked by applying the random-effects model or fixed. Besides, in order to seek out heterogeneity, the Galbraith plot analysis would be performed to determine whether outliers were the heterogeneity’s principal reason^21, 22^. Sensitivity analyses were conducted to see if there were any substantial changes in the results; if not, the results were more credible, and conversely, conclusions should be drawn with extreme caution. The Begg and Egger tests were used to evaluate the publication bias of the articles, *P* < 0.05 indicated that there was publication bias.

## Results

### Study Selection

Based on the Preferred Reporting Item for Systematic Reviews and Meta-Analyses (PRISMA) principles^23^, the selection procedure for the study was described in **Fig. 1**. A total of 258 articles were retrieved from the databases, of which 66 were excluded as duplicates. By reading the title and abstract of the remaining articles, 159 articles that did not meet the inclusion criteria were excluded. Then, 33 articles in total were taken for full-text review. 21 of the remaining articles were cut out because there weren’t sufficient information in them (n = 9), there were duplications (n = 2), and they were inconsistent with the subject matter (n = 10). 12 quality-appraised articles eventually became a portion of our meta-analysis^3, 16, 24–33^.

**Fig. 1.**
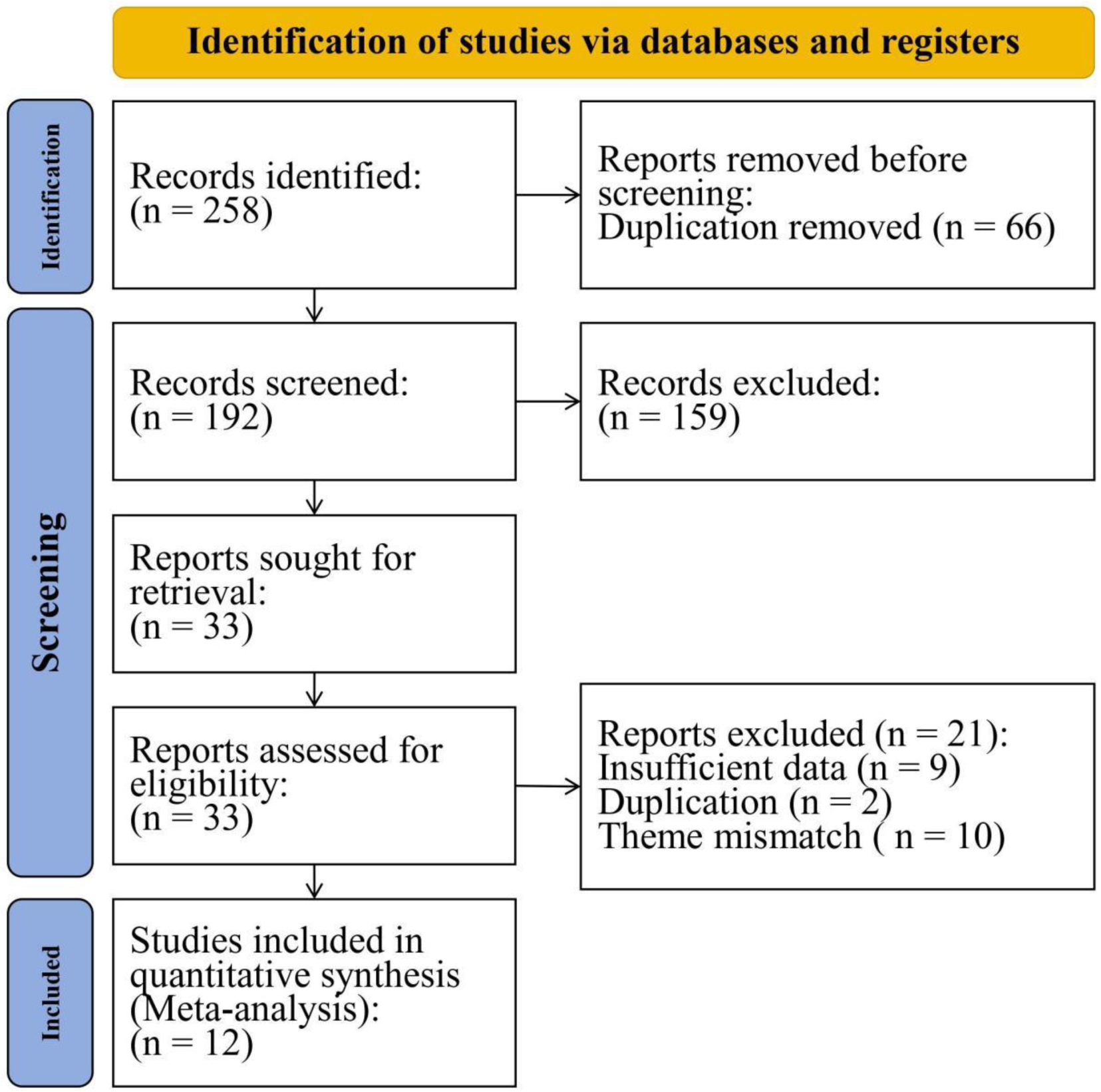
Flow diagram of study retrieval and screening

### Study characteristics

The included studies’ characteristics were showed in **Table 1**. All 12 studies included observational experiments, of which 11^3, 16, 25–33^ and 1^24^ were retrospective and prospective articles, respectively. Participants in the papers were from China; six of the papers were Chinese^28–33^ and six were English^3, 16, 24–27^. The subjects were 4708 patients with AIS, including 721 patients with HT and 3987 patients without HT. In these articles, the interventions involved were IVT (n = 7)^16, 26, 28, 30–33^ and EVT (n = 4)^24, 25, 27, 29^, respectively, and one of them did not limit IVT or EVT^3^. There were 8 articles believed that patients without HT had significantly higher UA degrees compared to those with HT^3, 16, 26–28, 30–32^. Several articles have enriched this conclusion: one of the articles^16^ clearly stated that no matter what types of HT, the UA degrees were lower than that of the non-HT group; another article^26^ drew the further conclusion that lower UA degrees were an independent risk factor for HT in large artery atherosclerosis stroke (LAA) patients or cardioembolism (CE) patients; and another paper^16^ indicated that ascending UA degrees may bring good outcomes compared to poor outcomes in AIS patients (345.67 ± 103.55 vs. 336.95 ± 95.5 μmol/L, *P* = 0.509). In addition, the best cutoff UA degrees for HT from 4 articles^16, 28, 30, 32^ were different: 218.5μmol/L, 284.00μmol/L, 364.5μmol/, 471.71μmol/L, respectively. And one paper’ s discussion^16^ showed that when the patient’ s UA degrees were lower than 218.5μmol/L or higher than 404.76μmol/L, the risk of HT may increase. On the contrary, one paper^25^ revealed that elevated UA degrees were not only risk factors but also predictors of SICH after EVT in patients. The data from 3 articles^24, 29, 33^ showed that the UA degrees in the HT group were higher than those in the non-HT group, but there were no significant differences (*P* > 0.05); one^29^ thought that hyperuricemia is a risk factor for AIS patients; and another^24^ showed that baseline high UA degrees may predict a better 90-day functional outcome. All studies had clearly identified diagnostic methods for AIS and HT

**Table 1.** Characteristics of the included studies.

| First name, year | Country Language | Design | N<br>M/F | Age<br>( Mean±SD ) | Therapy | HT types | HT timepoint | SUA timepoint | SUA cutoff<br>(μmol/L) | Segmentation of<br>SUA levels<br>(μmol/L) | Case |  | SUA (μmol/L) |  | Association Between SUA and<br>HT | NOS |
| --- | --- | --- | --- | --- | --- | --- | --- | --- | --- | --- | --- | --- | --- | --- | --- | --- |
|  |  |  |  |  |  |  |  |  |  |  | HT | non-HT | HT | non-HT |  |  |
| Shang<br>2023 | China<br>Chinese | Re | 100<br>61/39 | HT : 64.31±5.89<br>nHT : 66.19±6.3<br>65.81±6.24 | IVT | HT | 24 h after IVT | on admission | 471.71 | NR | 20 | 80 | 390.21 ± 102.16 | 471.32 ± 115.67 | positive | 4 |
| Zhang<br>2021 | China<br>Chinese | Re | 40<br>28/12 | 70.5±6.5 | IVT | HT | 24 h after IVT | on admission | NR | NR | 11 | 29 | 294.7±37.4 | 348.5±63.5 | positive | 5 |
| Chen<br>2021 | China<br>Chinese | Re | 258<br>147/111 | 61.37±12.36 | EVT | HT | 12h, 24h, 48h<br>after EVT | on admission | NR | NR | 62 | 196 | 311.25 ± 64.03 | 309.89 ± 57.16 | No significant association | 6 |
| Sun<br>2021 | China<br>Chinese | Re | 228<br>167/61 | 69.32±7.17 | IVT | HT | 24 h after IVT | on admission | 284.00 | NR | 30 | 198 | 281.20±77.79 | 340.63±78.95 | positive | 6 |
| Chen<br>2020 | China<br>Chinese | Re | 173<br>102/71 | 66.38±8.72 | IVT | HI<br>PH | 24 h after IVT | on admission | > 364.5 | NR | 46 | 127 | 324.89±70.43 | 383.08±89.21 | positive | 8 |
| Wei<br>2017 | China<br>Chinese | Re | 67<br>45/22 | HT : 73.25±8.87<br>nHT : 66.88±10.28<br>68.40±10.27 | IVT | HT | 72 h after IVT | on admission | NR | NR | 16 | 51 | 328.29 ± 49.48 | 306.86 ± 63.70 | No significant association | 4 |
| Tian,Y<br>2022 | China<br>English | Re | 727<br>509/218 | 64 | IVT | HI1 ; HI2 ;<br>PH1 ; PH2 | 7 days after<br>admission | 48 h after IVT | 218.50 | Q1 (<240.6)<br>Q2 (240.6–302)<br>Q3 (302–360)<br>Q4 (>360) | 112 | 615 | 253.65±97.75 | 315.97±96.42 | positive | 8 |
| Bai,H<br>2022 | China<br>English | Pro | 780<br>517/263 | 64.40±12.14 | EVT | sICH | 24 h after EVT | 24h after EVT | NR | Q1 (<245.85)<br>Q2 (245.85–297.45)<br>Q3 (297.45–364.95)<br>Q4 (≥364.95) | 47 | 733 | 319.72 ± 120.67 | 309.39 ± 94.51 | No significant association | 8 |
| Yang,C<br>2020 | China<br>English | Re | 247<br>177/70 | 63.2±12.4 | IVT | HI1 ; HI2 ;<br>PH1 ; PH2 ;<br>rPH | 72 h after IVT | at 6 AM the day<br>after admission. | NR | NR | 62 | 185 | 317.61±87.45 | 346.62±97.25 | positive;<br>No significance in multivariate<br>logistic regression analysis | 7 |
| Chen,Z<br>2020 | China<br>English | Re | 247<br>180/67 | 63.07±12.60 | EVT | HT<br>sICH | 72 h after<br>admission | at 6 AM on the<br>following day | NR | Q1 (<271)<br>Q2 (271–338)<br>Q3 (339–403)<br>Q4 (≥404) | 92 | 155 | 322.60±94.49 | 350.25±99.28 | positive | 7 |
| Yuan,K<br>2020 | China<br>English | Re | 611<br>366/245 | 64.7±12.3 | EVT | sICH | NR | 24 h after<br>admission | NR | Q1 (<247.2)<br>Q2 (247.2–310.0)<br>Q3 (310.0–380.8)<br>Q4 (≥380.8) | 90 | 521 | 341.0(291.1–391.0)<br>341.04±75.27 | 302.0(238.5–376.7)<br>305.92±102.74 | higher SUA level is risk factor<br>of sICH | 8 |
| Song,Q<br>2019 | China<br>English | Re | 1230<br>781/449 | 64.1±14.5 | IVT or<br>EVT | HI, PH ;<br>sHT | 7 days after<br>admission | 24 h after<br>admission | NR | Q1 (≤ 292.0)<br>Q2 (292.1–377.0)<br>Q3 (≥ 377.1) | 133 | 1097 | 272.1±70.9 | 350.9±105.6 | positive | 8 |

Re retrospective; Pro prospective; N number of patients; M/F male/female; HT hemorrhage transformation; non-HT non-hemorrhage transformation; SD standard deviation; IVT intravenous thrombolysis; EVT endovascular thrombectomy; HI hemorrhagic infarction;
PH parenchymal hematoma; rPH remote parenchymal hematoma; sICH symptomatic intracranial hemorrhage; sHT symptomatic hemorrhagic transformation; NR not reported; SUA uric acid; Q quartile of uric acid

### Study Quality

The quality evaluation was carried out by applying the NOS standards illustrated in **Table 1**. The NOS results indicated better quality in the included publications, including 7 studies of good quality and 5 papers of suboptimal quality.

### Meta-analysis results

The resulting pooled SMDs suggested that, compared with higher UA degrees, lower UA degrees were associated with HT after AIS (SMD = -0.34, 95% CI = -0.60–-0.08, *P* = 0.010) (**Fig. 2**). However, the high degree of heterogeneity (I^2^ = 89.0%, *P* < 0.001) was observed; thus, the random-effects model was used to pooled SMDs.

**Fig. 2.**
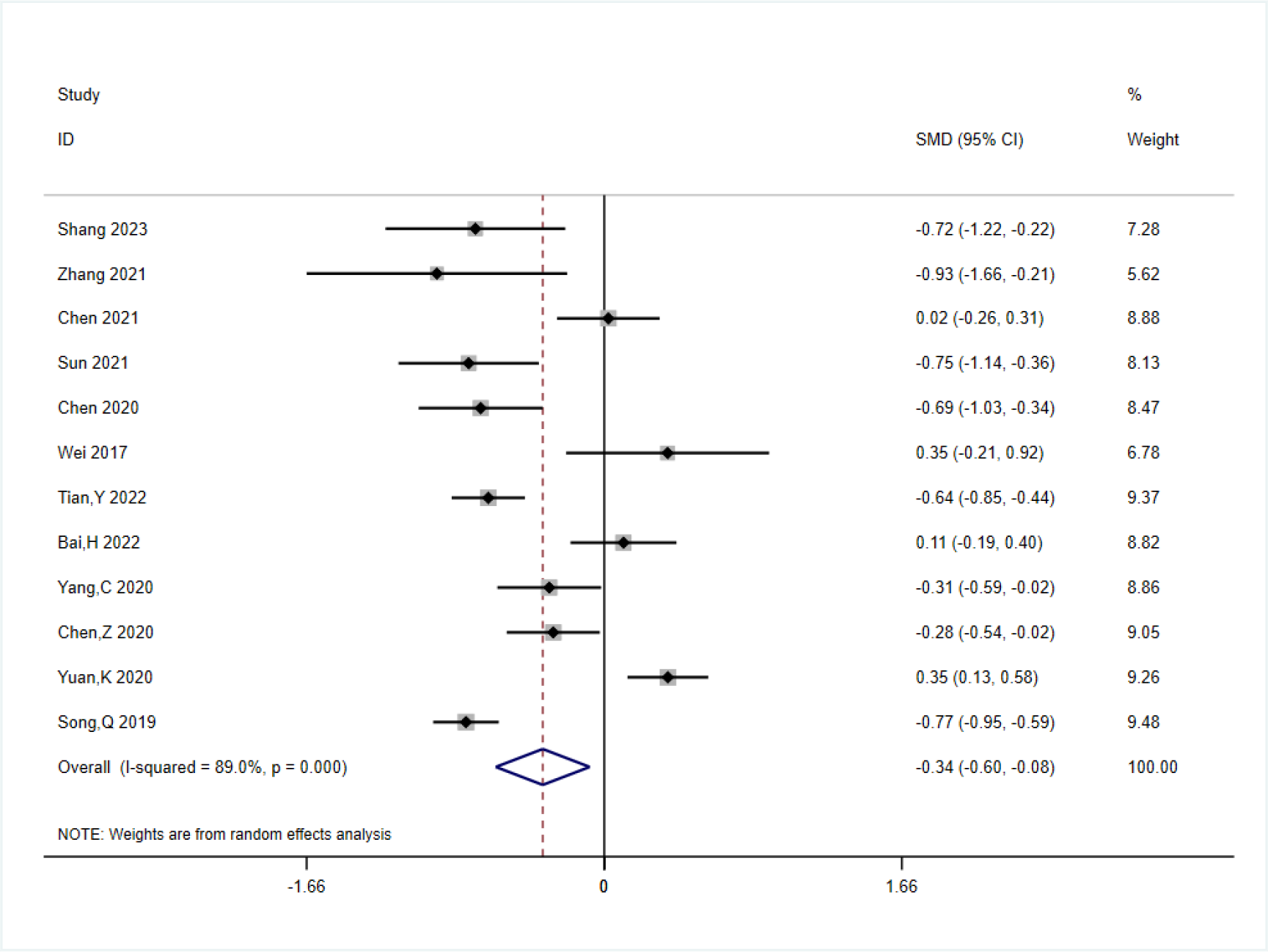
Lower UA degrees were linked with higher HT after AIS.

### Post-outlier analysis

There was shown to be considerable heterogeneity among the trials (I^2^ = 89.0%, *P* < 0.001), which prompted an investigation using sensitivity analysis (**Fig. 3**) and the Galbraith plot (**Fig. 4**). Sensitivity analyses tested the robustness of the results of the Meta-analysis by excluding one study in turn, and showed that the including 12 studies obtained consistent results in the pooled SMDs analysis. Applying the Galbraith plot analysis, outliers were identified as potential sources of heterogeneity. As pointed out by the Galbraith plot, half of the studies fell outside of the regression line. Excluding the outlier studies and then conducting the forest plot analysis again showed that there was no heterogeneity in the remaining six studies.(I^2^ = 45.8%, *P* = 0.100) (**Fig. 5**). Presumably, the heterogeneity of the research was induced by those outlier studies. Next, the outlier analysis and fixed-effects model analysis outcomes illustrated that the resulting pooled SMDs (SMD = -0.487, 95% CI = -0.632–-0.342, P = 0.000) still suggest that the UA degrees in the HT group were lower compared with the non-HT group.

**Fig. 3.**
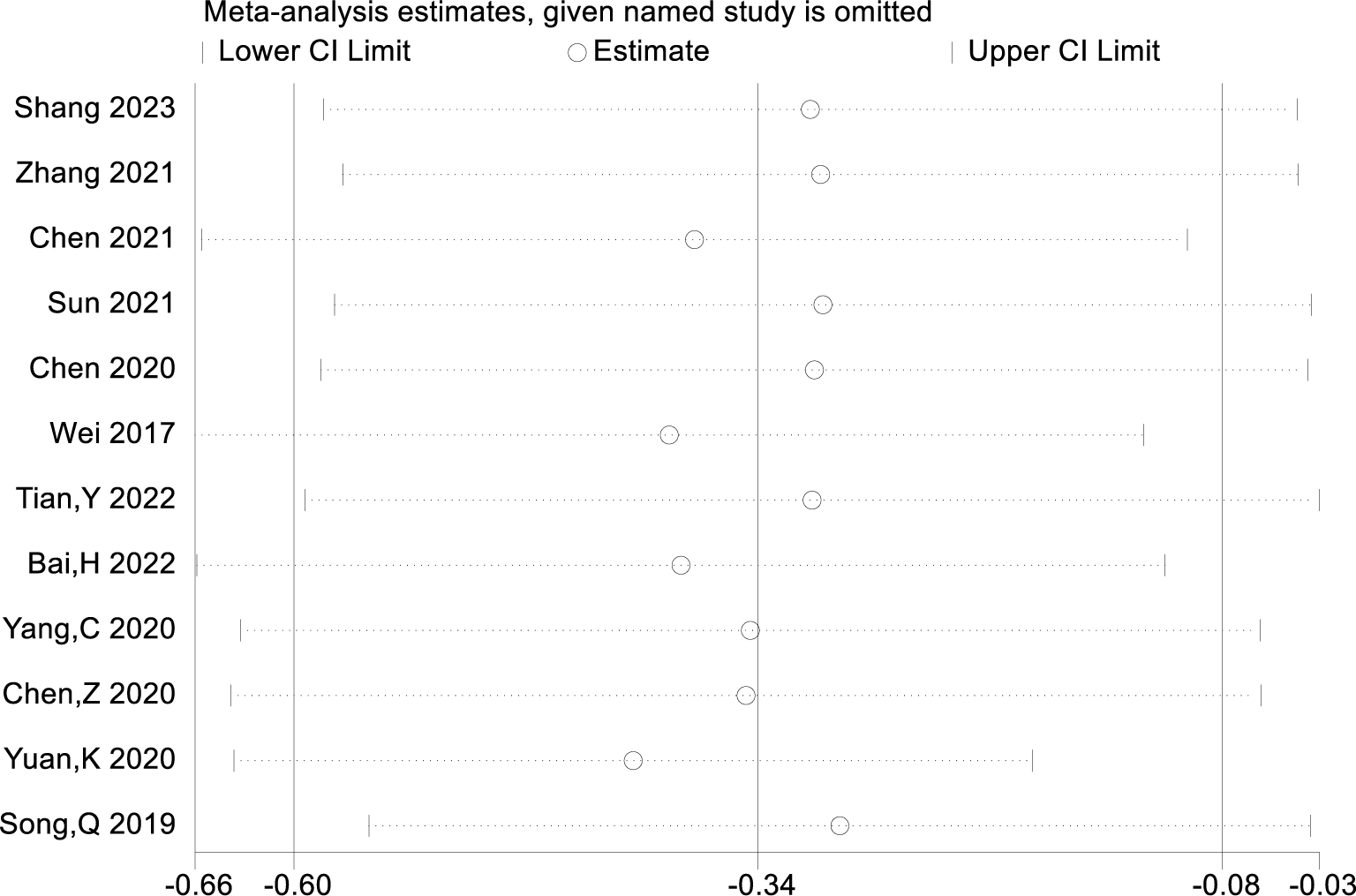
The provided study was taken away from the forest plot in the current meta-analysis’s sensitivity analysis.

**Fig. 4.**
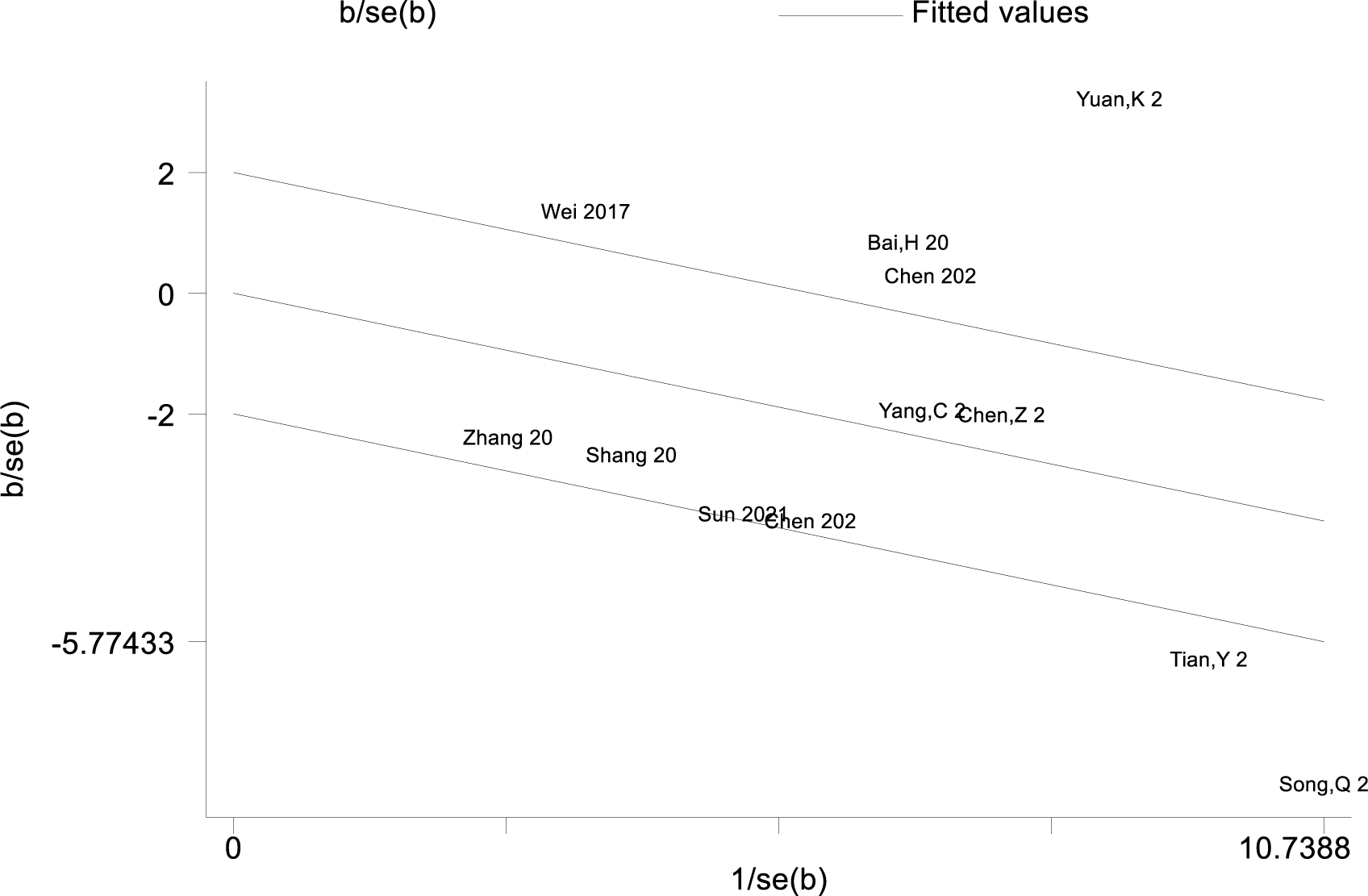
Galbraith plot analysis for outlier studies.

**Fig. 5.**
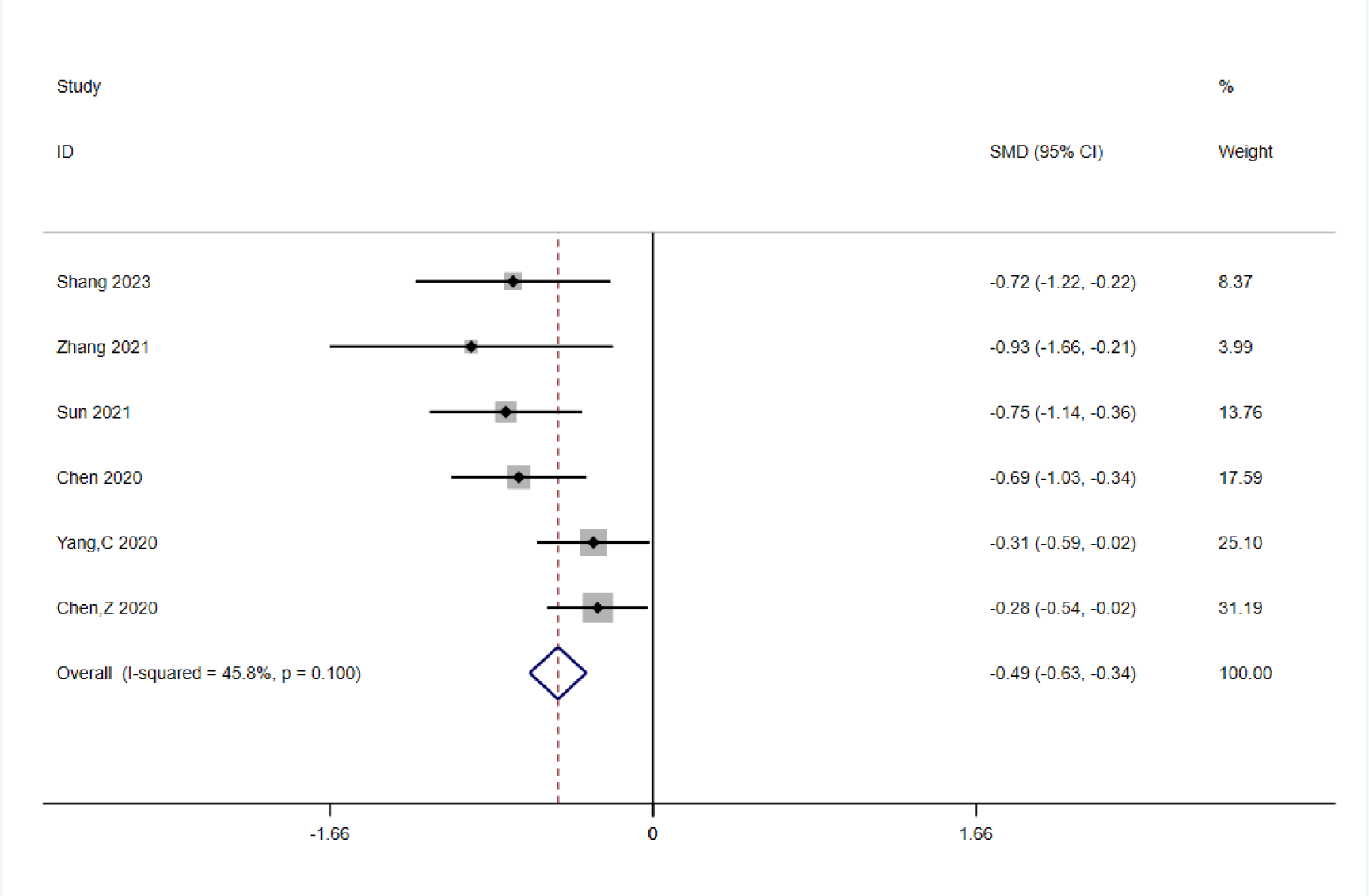
Forest plot analysis for the post-outlier outcomes indicated the UA degrees in the HT group were lower compared with the non-HT group.

### Publication bias

Begg’s funnel plot and Egger’s test of the 12 articles were used to evaluate the possible publication bias. Although the visual Begg’s funnel plot (**Fig. 6a**) was not completely symmetrical, the Egger’s test (**Fig. 6b**) with all *P* > 0.05 implied that there was no obvious publication bias in this meta-analysis, and the conclusion was more reliable.

**Fig. 6.**
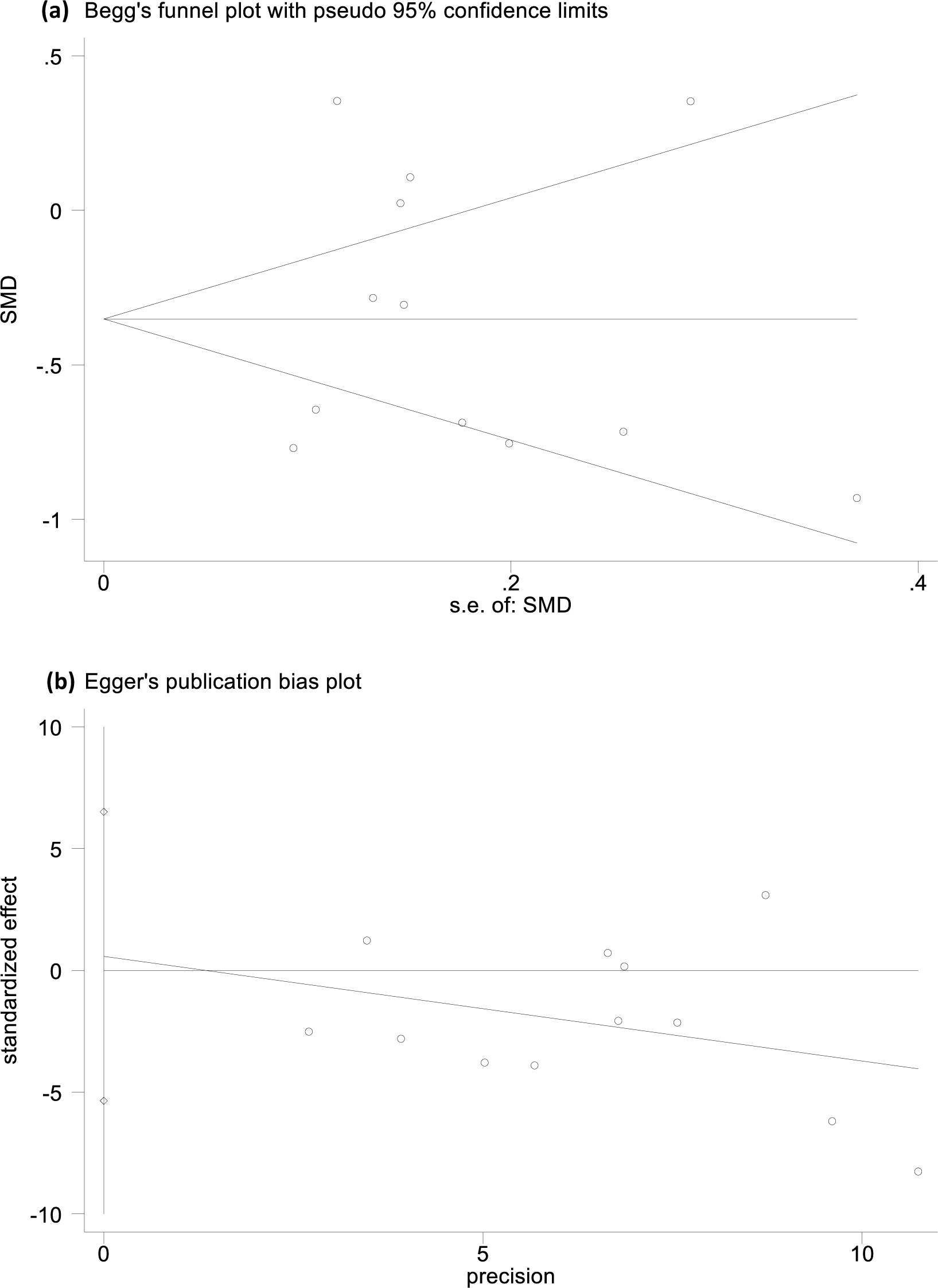
The funnel plot and Egger’s bias plot of publication bias in pooled SMDs analysis of all studies **(a and b)**.

## Discussion

In this meta-analysis, we identified 12 articles with 4,708 cases on the association between UA degrees and the risk of HT in AIS patients. These observations were made after accounting for the relevant covariates, implying that the lower UA degrees were independently associated with an increased risk of HT in patients with AIS. The articles we included were all from China, and the subjects included were patients with AIS who received IVT or EVT, with the primary outcome being HT. According to the neuroimaging, the patients were divided into HT group and non-HT group, while the MD or IQR values of UA in each group were collected. The forest plot of pooled SMDs suggested that, compared with high UA degrees, low UA degrees were associated with HT after AIS (SMD = -0.34, 95% CI = -0.60–-0.08, *P* = 0.010). Meanwhile, the high heterogeneity was discovered among the studies (I^2^ = 89.0%, *P* < 0.001). Initially, we performed subgroup analyses on language (Chinese or English), sample size of participants (≤250 or more), age (≤65 or older), treatment (IVT or EVT), HT time points, SUA time points, and adjusted OR (YES or NO) to find articles that may cause high heterogeneity, but there was still high heterogeneity among groups **(figures not shown)**. So we used sensitivity analysis and the Galbraith plot to process the heterogeneity. The reliability of the results of the Meta-analysis was demonstrated by the sensitivity analysis by leaving out one study consistently. The Galbraith plot analysis identified six studies as potential sources of heterogeneity. When deleting the outlier studies, the further forest plot of the remaining studies showed the I^2^ value decreased dramatically (I^2^ = 45.8%, *P* = 0.100), and the repeat pooled SMDs (SMD = -0.487, 95% CI = -0.632–-0.342, *P* = 0.000) still suggested that UA degrees are lower in the HT group. Moreover, Begg’s funnel plot and Egger’s test also showed no existence of publication bias in the Meta-analysis. The above analysis increased the credibility of our results.

More and more studies have confirmed that UA is a double-edged sword. It is unsurprisingly identified that UA accumulation is productive of the deposition of monosodium urate (MSU) crystals in the kidneys and joints, which can lead to kidney stones and gout. Epidemiological research has pointed out a correlation between elevated UA degrees and the happen of hypertension, cardiovascular and cerebrovascular events, insulin resistance, and diabetes mellitus. Nicotinamide adenine dinucleotide phosphate-oxidase (NADPH) is activated by UA, which performs the role of a pro-oxidant in the chemical microenvironment and can result in oxidative stress. UA can trigger inflammatory responses by releasing chemokines, inflammatory markers, and activating vasoconstrictive mediators, such as thromboxane, endothelin-1, and angiotensin II. Hyperuricemia reduces NO bioavailability, causing endothelial dysfunction, and urinary acid-lowering drugs can improve vascular endothelial function. In addition, UA also activate the renin-angiotensin system (RAS), causing vascular smooth muscle cell proliferation and arterial stiffness^34–36^. The numerous studies^13, 15^ have showed that UA has the possibility of independently predicting early death in AIS patients. Therefore, to a certain extent, high UA degrees may promote the occurrence of diseases and are not conducive to clinical prognosis. Four of the studies we included showed that UA degrees in the HT group were higher than those in the non-HT group, but only one result was significant (*P* < 0.05). The study suggested that adding UA degrees not only increased the risk of SICH after EVT in patients but also was a risk predictor of SICH^25^.

In the recent decade, UA, however, has gained important interest in stroke as a potential neuroprotective agent^37^, and more and more studies have focused on the anti-oxidant capacity of UA. As an endogenous extracellular antioxidant produced by purine metabolism, UA accounts for about 70 % of total antioxidant capacity^38^. It can inhibit the accumulation of reactive oxygen species and lipid peroxidation after exposure to glutamic acid or cyanide and scavenge free radicals produced by ischemia-reperfusion injury, and is considered to be neuroprotective. A comprehensive review and meta-analysis^39^ of the effects of UA on animal models of IS pointed out that elevated UA degrees after IS can reduce infarct size, improve BBB integrity, and enhance neurological function. Meanwhile, a recent clinical meta-analysis^10^ including 8,131 AIS patients and 10 eligible trials came to a similar conclusion: UA possessed a protective impact on neurological prognosis of AIS. Further, according to the neuroprotective effect of UA and the characteristics of significant decrease after the onset of the disease^34, 40^, it has tried to be used as a clinical treatment in stroke during the last decades^41^. A double-blind, randomized, vehicle-controlled study^42^ found that UA treatment can improve the prognosis of stroke by reducing the degree of MMP-9 to prevent oxidative stress. The URICO-ICTUS study^43^ pointed out that UA addition to thrombolytic treatment did not raise excellent outcomes in stroke patients in comparison with placebo but did not elevate safety worries.

For now, there is still an abundance of disagreement about the relationship between UA degrees and the risk of HT in AIS patients. This meta-analysis showed that high UA degrees in AIS patients reduced the likelihood of developing HT and were more favorable to the prognosis of AIS. The pathogenesis of HT after AIS was not yet clear, but relevant studies^3, 16, 44, 45^ believed that ischemia-reperfusion injury (IRI) will lead to free radical activation, resulting in amounts of reactive oxygen species (ROS) and reactive nitrogen species (RNS). ROS can promote glutamate release and calcium overload and result in neurotoxicity; it can also lead to cell necrosis and apoptosis by activating adhesion molecules, promoting leukocyte infiltration, and releasing a variety of cytokines. The high concentrations of RNS can induce matrix metalloproteinase (MMP) activation, mediate blood-brain barrier damage, expand infarct volume, and promote inflammation and apoptosis. When large number of free radicals are produced to cause oxidative stress, xanthine oxidase is activated and endogenous urea degrees increase, thereby inhibiting the activity of peroxynitrite and reducing neuronal damage. As a neuroprotectant, UA can also suppress the stiff inflammatory reaction that is brought on by ischemia, diminish vascular impairment and the breakdown of the blood-brain barrier, and decrease the infarct area by restraining oxygen’s creation of radicals on the blood vessel wall and reducing MMP-9 activity.

The analysis of articles revealed that UA was a protective factor for patients with AIS, and those with high UA degrees were less likely to develop HT, but the UA value was not the higher the better. In the enrolled studies, the best cutoff values of UA in HT patients were 218.5 μmol/L^16^, 284 μmol/L^30^, 364.5 μmol/L^32^, and 471.71 μmol/L^28^, respectively. The discussion of one of the articles clearly pointed out that the UA value should be between 218.5 μmol/L and 404.76 μmol/L, and lower or higher than that would lead to an increase in the incidence of HT^16^. Meanwhile, several other extraneous factors, including blood sample time, diverse biochemical analyzers, individual contrasts, and area of genesis, may have influence on the UA threshold value for HT. In our investigation, we thought that the existing studies could not give the nearest range value of UA, so large number of large-scale standardized clinical studies are needed in the future.

There are gender differences in UA degrees, and studies have confirmed that the average UA degree is significantly lower in female than in male due to estrogen^3, 46^. A sex-stratified analysis of one study we brought to a close demonstrated that^3^, UA degrees in male were significantly higher than those in female, and the incidence of HT was lower than that in female, which corresponded with earlier discoveries, as anticipated. This also side-steps our results that UA was a protective factor for HT post-AIS.

We were not able to reduce the substantial heterogeneity in our study by excluding individual studies. The studies themselves, the study design, the parameter-assessing manner, the exceedingly varying duration of follow-up, and so forth could be potential causes of high heterogeneity. Due to the high heterogeneity, the results were not as robust as expected and should be treated with caution, but the preliminary findings were still noteworthy.

Several limitations needed attention in our study. First, this study had high heterogeneity. Practicing the Galbraith plot analysis, we were able to discover six studies that could be the sources of heterogeneity. Due to no report or incomplete information in the literature, it was not feasible to identify the reasons for the high heterogeneity induced by these six articles. Second, there were insufficient data to conduct subgroup analyses that included AIS type, HT types and gender. Further clinical studies targeting relevant subgroups could be conducted in the future. Third, each included study obtained only one measurement of UA, which did not take into account the dynamic fluctuation of UA in the course of AIS. Therefore, more studies are needed to measure UA values multiple times to dynamically monitor the relationship between UA and HT. Eventually, all the studies were conducted in China, and the applicability of our results in other regions was limited.

## Conclusion

To a certain extent, the meta-analysis found that UA played a neuroprotective role in AIS patients and lower UA degrees in AIS patients may be a risk factor for HT, which provided some theoretical basis for the following related studies. At the same time, some problems also need to be further studied in the future, such as UA dynamic degree, UA optimal range, some subgroups based on AIS and HT, etc.

## CRediT authorship contribution statement

**Ying Qian**: wrote the original manuscript, **Na Li**: collected data and revised articles, **Yuanyuan Li and Chenxi Tao**: extracted and analyzed the outcomes, **Guoxia Zhang and Fan Yang**: assessed the risk of bias, **Zhenhong Liu and Hongrui Zhang**: checked and modified the language and grammar, **Yong-hong Gao**: critically revised, scientifically reviewed and edited.

## Data Availability

The data on which the study is based were accessed from a repository and are available for downloading through the following link.

https://www.cnki.net/

https://pubmed.ncbi.nlm.nih.gov/

## Acknowledgements

This study was supported by the National Natural Science Foundation of China (8227151735).

## References

1. Chiquete E, Ruiz-Sandoval JL, Murillo-Bonilla LM, Arauz A, Orozco-Valera DR, Ochoa-Guzmán A, et al. Serum uric acid and outcome after acute ischemic stroke: Premier study. *Cerebrovascular diseases (Basel*, Switzerland). 2013;35:168–174

2. Zhang W, Cheng Z, Fu F, Zhan Z. Serum uric acid and prognosis in acute ischemic stroke: A dose-response meta-analysis of cohort studies. Frontiers in aging neuroscience. 2023;15:1223015

3. Song Q, Wang Y, Cheng Y, Liu J, Wei C, Liu M. Serum uric acid and risk of hemorrhagic transformation in patients with acute ischemic stroke. Journal of molecular neuroscience : MN. 2020;70:94–101

4. Brouns R, Wauters A, Van De Vijver G, De Surgeloose D, Sheorajpanday R, De Deyn PP. Decrease in uric acid in acute ischemic stroke correlates with stroke severity, evolution and outcome. Clinical chemistry and laboratory medicine. 2010;48:383–390

5. Chamorro A, Obach V, Cervera A, Revilla M, Deulofeu R, Aponte JH. Prognostic significance of uric acid serum concentration in patients with acute ischemic stroke. Stroke. 2002;33:1048–1052

6. Liu C, Xie J, Sun S, Li H, Li T, Jiang C, et al. Hemorrhagic transformation after tissue plasminogen activator treatment in acute ischemic stroke. Cellular and molecular neurobiology. 2022;42:621–646

7. Ma G, Pan Z, Kong L, Du G. Neuroinflammation in hemorrhagic transformation after tissue plasminogen activator thrombolysis: Potential mechanisms, targets, therapeutic drugs and biomarkers. International immunopharmacology. 2021;90:107216

8. Kanazawa M, Takahashi T, Nishizawa M, Shimohata T. Therapeutic strategies to attenuate hemorrhagic transformation after tissue plasminogen activator treatment for acute ischemic stroke. Journal of atherosclerosis and thrombosis. 2017;24:240–253

9. Arba F, Rinaldi C, Caimano D, Vit F, Busto G, Fainardi E. Blood-brain barrier disruption and hemorrhagic transformation in acute ischemic stroke: Systematic review and meta-analysis. Frontiers in neurology. 2020;11:594613

10. Wang Z, Lin Y, Liu Y, Chen Y, Wang B, Li C, et al. Serum uric acid levels and outcomes after acute ischemic stroke. Molecular neurobiology. 2016;53:1753–1759

11. Dong Y, Shi H, Chen X, Fu K, Li J, Chen H, et al. Serum uric acid and risk of stroke: A dose-response meta-analysis. Journal of clinical biochemistry and nutrition. 2021;68:221–227

12. Tariq MA, Shamim SA, Rana KF, Saeed A, Malik BH. Serum uric acid -risk factor for acute ischemic stroke and poor outcomes. Cureus. 2019;11:e6007

13. Chen JH, Chuang SY, Chen HJ, Yeh WT, Pan WH. Serum uric acid level as an independent risk factor for all-cause, cardiovascular, and ischemic stroke mortality: A chinese cohort study. Arthritis and rheumatism. 2009;61:225–232

14. Weir CJ, Muir SW, Walters MR, Lees KR. Serum urate as an independent predictor of poor outcome and future vascular events after acute stroke. Stroke. 2003;34:1951–1956

15. Karagiannis A, Mikhailidis DP, Tziomalos K, Sileli M, Savvatianos S, Kakafika A, et al. Serum uric acid as an independent predictor of early death after acute stroke. Circulation journal : official journal of the Japanese Circulation Society. 2007;71:1120–1127

16. Tian Y, Xie Q, You J, Yang S, Zhao H, Song Y. Lower uric acid level may be associated with hemorrhagic transformation after intravenous thrombolysis. Neurological sciences : official journal of the Italian Neurological Society and of the Italian Society of Clinical Neurophysiology. 2022;43:3113–3120

17. Lei Z, Cai J, Hong H, Wang Y. Serum uric acid level and outcome of patients with ischemic stroke: A systematic review and meta-analysis. The neurologist. 2019;24:121–131

18. Stang A. Critical evaluation of the newcastle-ottawa scale for the assessment of the quality of nonrandomized studies in meta-analyses. European journal of epidemiology. 2010;25:603–605

19. Wan X, Wang W, Liu J, Tong T. Estimating the sample mean and standard deviation from the sample size, median, range and/or interquartile range. BMC medical research methodology. 2014;14:135

20. Luo D, Wan X, Liu J, Tong T. Optimally estimating the sample mean from the sample size, median, mid-range, and/or mid-quartile range. Statistical methods in medical research. 2018;27:1785–1805

21. Wang L, Deng ZR, Zu MD, Zhang J, Wang Y. The comorbid relationship between migraine and asthma: A systematic review and meta-analysis of population-based studies. Frontiers in medicine. 2020;7:609528

22. Arceo ES, Dizon GA, Tiongco REG. Serum cystatin c as an early marker of nephropathy among type 2 diabetics: A meta-analysis. Diabetes & metabolic syndrome. 2019;13:3093–3097

23. Moher D, Shamseer L, Clarke M, Ghersi D, Liberati A, Petticrew M, et al. Preferred reporting items for systematic review and meta-analysis protocols (prisma-p) 2015 statement. Systematic reviews. 2015;4:1

24. Bai H, Nie X, Leng X, Wang D, Pan Y, Yan H, et al. Increased serum uric acid level is associated with better outcome after endovascular treatment for acute ischemic stroke-a prospective cohort study. Annals of translational medicine. 2022;10:1111

25. Yuan K, Zhang X, Chen J, Li S, Yang D, Xie Y, et al. Uric acid level and risk of symptomatic intracranial haemorrhage in ischaemic stroke treated with endovascular treatment. European journal of neurology. 2020;27:1048–1055

26. Yang C, Zhang J, Liu C, Xing Y. Comparison of the risk factors of hemorrhagic transformation between large artery atherosclerosis stroke and cardioembolism after intravenous thrombolysis. Clinical neurology and neurosurgery. 2020;196:106032

27. Chen Z, Chen H, Zhang Y, He Y, Su Y. Lower uric acid level may be associated with hemorrhagic transformation but not functional outcomes in patients with anterior circulation acute ischemic stroke undergoing endovascular thrombectomy. Metabolic brain disease. 2020;35:1157–1164

28. Fenglan. S, Jianpu. L. Value of serum uric acid and homocysteine in predicting the transformation of hemorrhage after intravenous thrombolysis in patients with acute ischemic stroke. Clinical Medicine & Engineering. 2023;30:1023–1024. (in Chinese)

29. Yingdao. C, Haining. L, Yuying. L, al. e. Distribution characteristics of risk factors for acute ischemic stroke and influencing factors of hemorrhagic transformation after stent thrombectomy. Chinese Journal of Stereotactic and Functional Neurosurgery. 2021;34:262–268

30. Ruonan. S, Yuanzheng. Z, Yinghui. Z, Zhiwen. C, Shanshan. L, Heping. Y, et al. Value of serum calcium and ua levels before thrombolysis for predicting ht in ais patients after intravenous thrombolysis. Chinese Journal of Geriatric Heart Brain and Vessel Diseases. 2021;23:617–620. (in Chinese)

31. Hui. Z, Xiaohong. C, Ying. L. Clinical observation of intrusive bleeding during intravenous rt-pa thrombolysis in patients with acute ischemic stroke. Chinese General Practice. 2021;24:85–88. (in Chinese)

32. Chen Y, Zhang Q, You N, Wang L. [analysis of influencing factors of neurological function recovery and cerebral hemorrhage transformation after intravenous thrombolysis in patients with acute ischemic stroke]. Zhonghua wei zhong bing ji jiu yi xue. 2020;32:1340–1345

33. Xiaofan. W, Chunhui. C, Changyun. L, al e. Analysis of risk factors related to hemorrhagic transformation after thrombolytic therapy in patients with acute ischemic stroke. Chinese Journal of Clinical Rational Drug Use. 2017;10:15–16+18. (in Chinese)

34. Lim SS, Yang YL, Chen SC, Wu CH, Huang SS, Chan WL, et al. Association of variability in uric acid and future clinical outcomes of patient with coronary artery disease undergoing percutaneous coronary intervention. Atherosclerosis. 2020;297:40–46

35. Sun J, Lv X, Gao X, Chen Z, Wei D, Ling Y, et al. The association between serum uric acid level and the risk of cognitive impairment after ischemic stroke. Neuroscience letters. 2020;734:135098

36. Li M, Huang Y, Lin H, Chen Y. Association of uric acid with stenosis of intracranial and extracranial arteries in elderly patients with cerebral infarction. Neurological sciences : official journal of the Italian Neurological Society and of the Italian Society of Clinical Neurophysiology. 2019;40:957–961

37. Becker BF. Towards the physiological function of uric acid. Free radical biology & medicine. 1993;14:615–631

38. Mármol F, Sanchez J, Martínez-Pinteño A. Effects of uric acid on oxidative and nitrosative stress and other related parameters in sh-sy5y human neuroblastoma cells. Prostaglandins, leukotrienes, and essential fatty acids. 2021;165:102237

39. Aliena-Valero A, Baixauli-Martín J, Castelló-Ruiz M, Torregrosa G, Hervás D, Salom JB. Effect of uric acid in animal models of ischemic stroke: A systematic review and meta-analysis. Journal of cerebral blood flow and metabolism : official journal of the International Society of Cerebral Blood Flow and Metabolism. 2021;41:707–722

40. Fernández-Gajardo R, Matamala JM, Gutiérrez R, Lozano P, Cortés-Fuentes IA, Sotomayor CG, et al. Relationship between infarct size and serum uric acid levels during the acute phase of stroke. PloS one. 2019;14:e0219402

41. Llull L, Amaro S, Chamorro Á. Administration of uric acid in the emergency treatment of acute ischemic stroke. Current neurology and neuroscience reports. 2016;16:4

42. Amaro S, Llull L, Renú A, Laredo C, Perez B, Vila E, et al. Uric acid improves glucose-driven oxidative stress in human ischemic stroke. Annals of neurology. 2015;77:775–783

43. Chamorro A, Amaro S, Castellanos M, Segura T, Arenillas J, Martí-Fábregas J, et al. Safety and efficacy of uric acid in patients with acute stroke (urico-ictus): A randomised, double-blind phase 213/3 trial. Lancet Neurol. 2014;13:453–460

44. Cheng Z, Zhan Z, Fu Y, Zhang WY, Xia L, Xu T, et al. U-shaped association between serum uric acid and hemorrhagic transformation after intravenous thrombolysis. Current neurovascular research. 2022;19:150–159

45. Aliena-Valero A, Rius-Pérez S, Baixauli-Martín J, Torregrosa G, Chamorro Á, Pérez S, et al. Uric acid neuroprotection associated to il-6/stat3 signaling pathway activation in rat ischemic stroke. Molecular neurobiology. 2021;58:408–423

46. Tang Y, Liu MS, Fu C, Li GQ. Sex-dependent association analysis between serum uric acid and spontaneous hemorrhagic transformation in patients with ischemic stroke. Frontiers in neurology. 2023;14

